# A random forest model for forecasting regional COVID-19 cases utilizing reproduction number estimates and demographic data

**DOI:** 10.1101/2021.05.23.21257689

**Authors:** Joseph Galasso, Duy M. Cao, Robert Hochberg

**Affiliations:** Department of Biology, #11, University of Dallas, Irving, TX 75062; Department of Computer Science, #134, University of Dallas, Irving, TX 75062; Department of Computer Science, #50, University of Dallas, Irving, TX 75062

**Keywords:** COVID-19, random forest, compartmental model, mobility, US county

## Abstract

During the COVID-19 pandemic, predicting case spikes at the local level is important for a precise, targeted public health response and is generally done with compartmental models. The performance of compartmental models is highly dependent on the accuracy of their assumptions about disease dynamics within a population; thus, such models are susceptible to human error, unexpected events, or unknown characteristics of a novel infectious agent like COVID-19. We present a relatively non-parametric random forest model that forecasts the number of COVID-19 cases at the U.S. county level. Its most prioritized training features are derived from easily accessible, standard epidemiological data (i.e., regional test positivity rate) and the effective reproduction number (*R*_*t*_) from compartmental models. A novel input training feature is case projections generated by aligning estimated effective reproduction number (pre-computed by COVIDActNow.org) with real time testing data until maximally correlated, helping our model fit better to the epidemic’s trajectory as ascertained by traditional models. Poor reliability of *R*_*t*_ is partially mitigated with dynamic population mobility and prevalence and mortality of non-COVID-19 diseases to gauge population disease susceptibility. The model was used to generate forecasts for 1, 2, 3, and 4 weeks into the future for each reference week within 11/01/2020 - 01/10/2021 for 3068 counties. Over this time period, it maintained a mean absolute error (MAE) of less than 300 weekly cases/100,000 and consistently outperformed or performed comparably with gold-standard compartmental models. Furthermore, it holds great potential in ensemble modeling due to its potential for a more expansive training feature set while maintaining good performance and limited resource utilization.

## 1 Introduction

The COVID-19 epidemic in the United States proved devastating economically as it is projected to cause 3.2 to 4.8 trillion USD in net U.S. GDP loss [1]. The epidemic has had a devastating toll on life, particularly among the elderly and members of ethnic minorities such as African Americans [2]. Throughout the COVID-19 pandemic, it has been necessary to forecast the progression of the pandemic at the U.S. county/county-equivalent (CCE) level so that local authorities can effectively implement public health measures such as social distancing or quarantines [3]. This need is particularly great due to findings that there is low reliability between state-wide and county-specific reported data, necessitating that the pandemic is tracked at the most granular level possible [4].

Forecasts have been traditionally done with compartmental models such as Susceptible-Exposed-Infectious-Recovered (SEIR) [5, 6]. These models segment the population into various compartments for stages in progression of the disease of interest. Transitions between these compartments represent epidemic dynamics. These models can be used to solve for the time-path of the *R*_*t*_, which is the estimated ratio of new infections caused by each currently infected individual, over the course of an epidemic; this in turn can be used to forecast future epidemic progression [6]. However, these models involve numerous assumptions in their design about disease spread dynamics and their interpretability and usability is limited by the quality of these assumptions [5, 6, 7].

Thus, relatively non-parametric deep or machine-learning models, such as Random Forests (RFs), are attractive alternatives to compartmental models, as they avoid assumptions about the distribution of input data and generate forecasts based on observed empirical trends in this data [8, 9]. In addition to being non-parametric, RF-regressors are highly effective at extracting non-linear relationships from input data while being time efficient [10, 11]. First described by Breiman, RF-regressors are ensemble models of regression trees each trained on different subsets of input data, reducing the variance of predictors and minimizing overfitting [12]. In addition, RFs enable relatively easy estimations of variable importance [13], which can serve as an assessment of model performance. RFs have been successfully utilized in prior studies for predicting diarrheal infections, Dengue, H5N1 influenza, and West Nile virus [10, 14, 15, 16, 17]. They also have been applied for short-term forecasting of COVID-19 infections by Ribeiro et al. [11].

We similarly apply an RF to project COVID-19 infections at the more granular CCE level up to 4 weeks in the future. Unlike prior RF epidemiology forecasting models, we use a unique and extensive combination of features including population health and mobility, demographic, and COVID-19 testing data. We develop a novel method of computing case and *R*_*t*_ forecasts with an algorithm that is based on our observation that COVID-19 cases and SEIR-generated *R*_*t*_ have a similar trajectory over the course of an epidemic, but *R*_*t*_ lags behind cases, making it predictive of cases in the lag period. These forecasts are then added as a training feature for our RF model, which produces the final forecasts.

## 2 Methods

### 2.1 Data Acquisition

Our time-series datasets begin on 03/31/2020 for 3068 U.S. CCEs with complete, consistent data that could be processed without error in our downstream pipelines. To normalize testing and case counts by population, these metrics were converted to incidence rate (IR) using the 2018 U.S. Census CCE-level population estimates in Eq. 1.

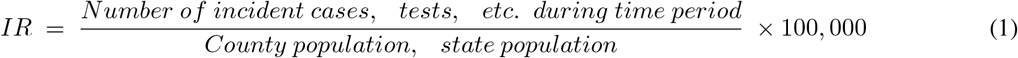

### 2.2 Generation of *R*_*t*_ and Case Alignment Forecast Features

For any given date in every U.S. CCE, we forecasted *R*_*t*_ and testing-normalized cases for 1, 2, 3, and 4 weeks into the future. First, a testing-normalized COVID-19 cases time-series was generated by dividing the “Weekly case increase” feature by the “Daily tests increase” feature (see Table 1). The normalized cases time-series and the “Daily estimated *R*_*t*_” feature were used to generate *R*_*t*_ and case-prediction features, as shown in Fig. 1. All linear regression models were implemented with the *linear*_*model*.*LinearRegression* module [28].

**Table 1:**
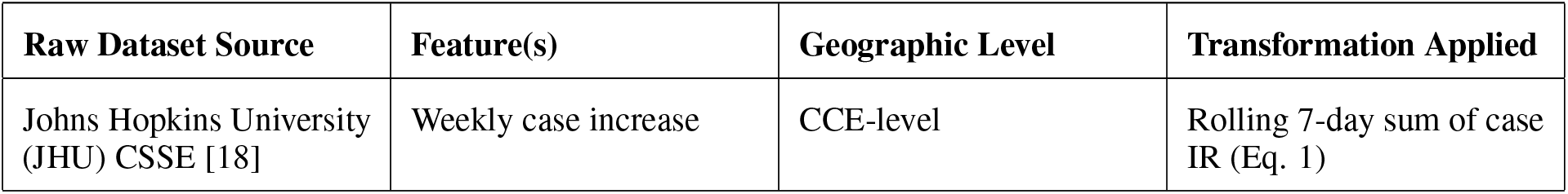
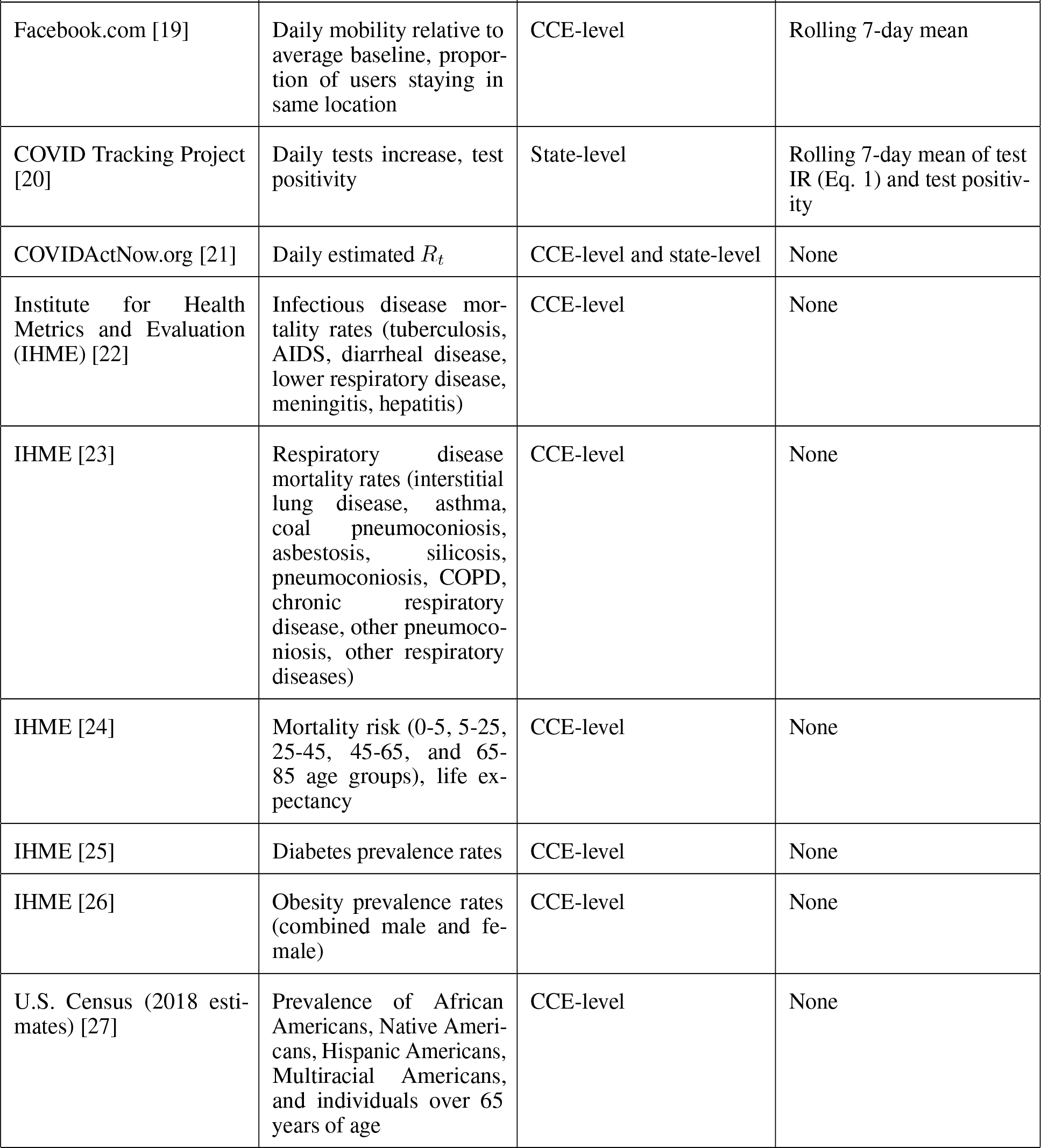
Raw Training Data Sources and Normalizations. Description of datasets, variables extracted, regional level, and applied normalization in the training dataset.

**Figure 1:**
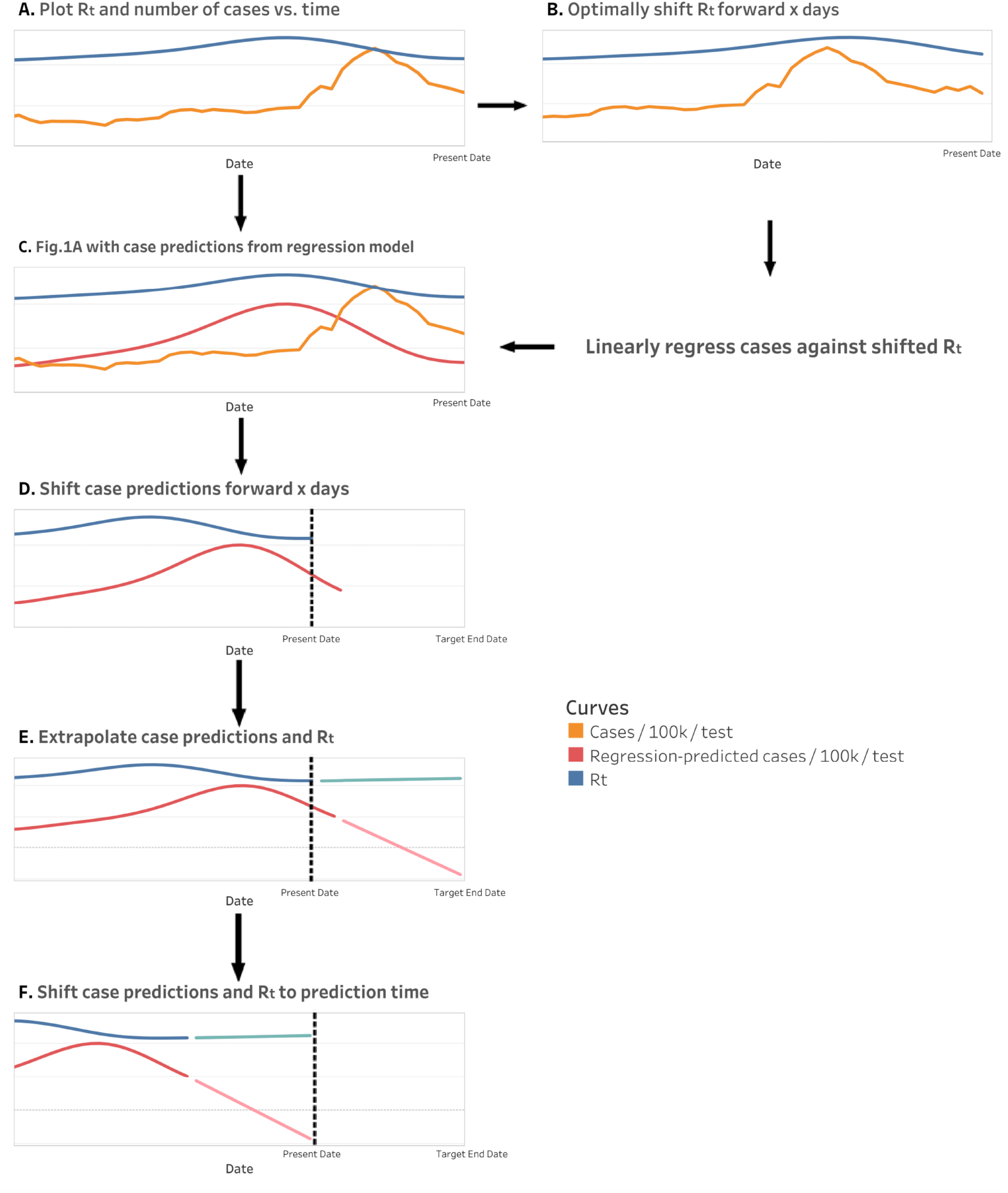
R_t_ and case prediction feature generation for a CCE. This procedure is repeated for 7-, 14-, 21-and 28-day forecasts. In Fig. 1A and Fig. 1B, *R*_*t*_ for the CCE and its state are both separately considered; whichever achieves the highest Pearson correlation of any forward shift x that is <50 days (i.e. optimal shift) is used for the regression model in Fig. 1B. The extrapolation in Fig. 1E is calculated by linear regression models trained on the last 14 defined values of each curve; curves are extrapolated to the target end date (i.e. 7, 14, 21, or 28 days in the future). For Fig. 1F, curves in prediction time have forecasted values relative to real time; thus, for 28-day forecasts, values are those forecasted 28 days into the future.

### 2.3 RF Training and Forecasting

The dataset used to train and validate the RF forecasting models includes all features in Table 1 for each U.S. CCE; however, “Weekly case increase” and “Daily estimated *R*_*t*_” features were replaced with the Fig. 1F features for 7-, 14-, 21-, and 28-day forecasts. Thus, the preliminary forecasts were used as features to generate the final forecasts. All features were normalized by removing their mean and scaling to unit variance. For each Sunday from 11/01/2020 to 01/10/2021, we trained and validated RF forecasting models via incremental learning. Thus, we filtered out feature data that occurs on or after the Sunday of interest, but later Sundays will have more feature data than earlier Sundays. Then, we randomly split the feature data for a given Sunday into a training subset and a validation subset with a 9:1 ratio, respectively. The training subset is used to train a RF regression model for each forecast timepoint (7, 14, 21, or 28 days into the future); the target outcome was the “Weekly case increase” from Table 1 shifted backwards to represent future cases on the forecast timepoint. The validation subset is used to ensure that trained RF models do not overfit the training subset. Finally, data for the Sunday of interest is input to the trained RF models to generate forecasts for each forecast timepoint.

RF regression models are ensemble machine learning algorithms that were first described by Breiman [12]. They create multiple regression trees trained on unique bootstrap samples of the training dataset with a random subset of the input features. The output of all trees is averaged to create the final projection. We used the Scikit-learn (version 0.23.2) implementation with default hyperparameters and 20 estimators [28].

### 2.4 Model Evaluation

The permutation importance of all features input into the RF models was calculated as described by Breiman as the decrease in mean squared error of the model when a feature of interest is randomly shuffled [12]. We used two metrics, MAE and R-squared (*R*^2^), to evaluate the accuracy of our model’s forecasts relative to actual case counts for both the training dataset and forecasts outside of the training dataset. These metrics are calculated as follows:

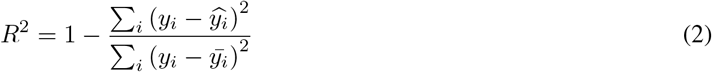

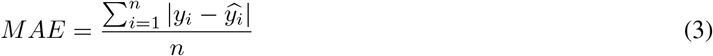

We also used these metrics to calculate the error for equivalent forecasts by the JHU Infectious Disease Dynamics group (IDD), the JHU Applied Physics Lab (APL), and One Quiet Night (OQN) forecasting models [29, 30, 31]. These were selected as they are parts of the U.S. Centers for Disease Control and Prevention (U.S. CDC) ensemble model, have relatively high performance, and forecast for a large breadth (2349) of U.S. counties along with the RF model [32].

## 3 Results and Discussion

### 3.1 Analysis of *R*_*t*_ and Case Alignment Forecasts

When the case time-series for a CCE was normalized by population and state-level testing data, it often showed a very similar shape to the CCE and/or state *R*_*t*_ time-series, as shown in Fig. 1. This should be expected, as both time-series indicate new infection load over the course of an epidemic, although *R*_*t*_ lags behind cases, which we suspect is because *R*_*t*_ represents infection load in the present moment, but these infections will not be detected via testing until much later due to viral incubation periods of 2-14 days and test result reporting delays [33]. In the 01/10/2021 dataset, the average optimal shift for the most highly correlated *R*_*t*_ time-series, whether state or CCE, was 34 days forward.

However, there were many CCEs where this relationship was weak. We observed that 1782 CCEs’ selected, optimally shifted *R*_*t*_ time-series have a Pearson correlation with the case time-series < 0.5. They also have a mean population density of 89 people/mi^2^, vs. the 273 people/mi^2^ mean over the entire set of 3068 CCEs. Thus, we attribute the low correlations to poor and/or inconsistent testing efforts and data quality in rural CCEs. Further supporting this is our observation that in CCEs where we select their state’s *R*_*t*_ time-series as opposed to their own, the mean population density is also relatively low at 210 people/mi^2^.

In concurrence with Omori et al., we found that normalization of the case time-series with testing data is critical to expose underlying changes in COVID-19 progression, as seen in Fig. 2 [34]. However, our approach is limited by use of state-level testing data as opposed to CCE-level testing data, which was inaccessible to us. However, state-level testing data has the advantage of including individuals who are not tested in their CCE of residence due to inequity in regional testing access.

**Figure 2:**
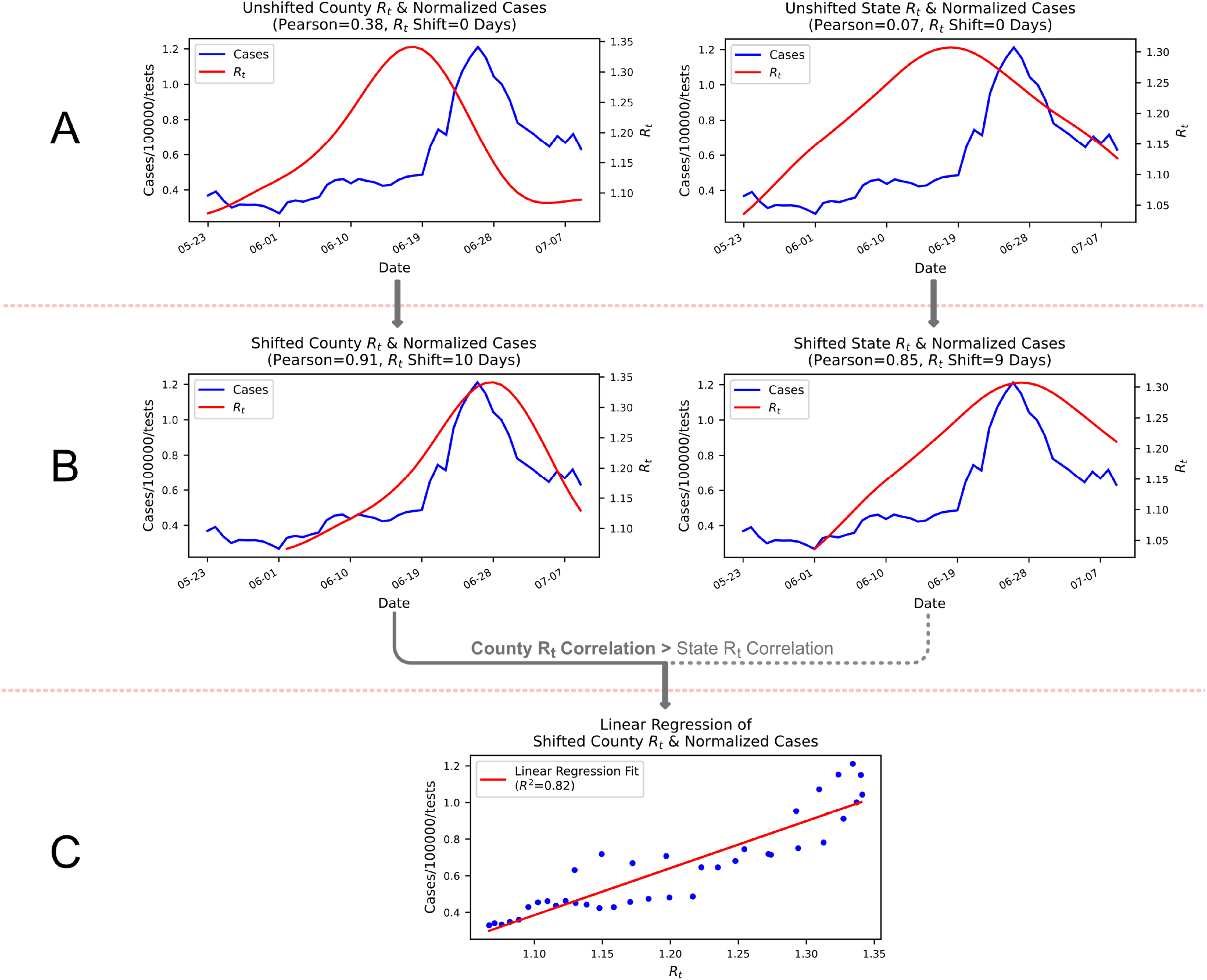
R_t_ Time-Series’ Lag Behind Case Time-Series Used to Forecast Cases in Lag Period. In Harris County, TX, when the CCE *R*_*t*_ time-series from 05/22/2020 to 07/10/2020 has a maximum Pearson correlation with the CCE’s testing and population normalized case time-series for the same period when shifted forward 10 days (B); also, this correlation is higher than that obtained by any shift of the state *R*_*t*_ time-series. Thus, the shifted CCE *R*_*t*_ time-series is linearly regressed against cases (C). This model is applied to unshifted CCE *R*_*t*_ time-series to generate a case-prediction time-series; the last 10 days of both these time-series are predictive of the next 10 days of cases beyond 07/10/2020.

### 3.2 RF Feature Prioritization and Training Performance

On average over the 11 epi weeks, RFs always prioritized population-normalized, state-level test numbers, test positivity, and case and *R*_*t*_ alignment forecasts as seen in Fig.3. This validates the model, as these features most directly measure COVID-19 progression. Importantly, of all 8 alignment forecasts, those for the desired forecast target were always prioritized highest. Conversely, demographic, population health, and population mobility features are relatively inconsequential; on average across Fig. 3A-D, these 46 features’ share of the total sum of all 56 features’ permutation scores is just 14.33%.

**Figure 3:**
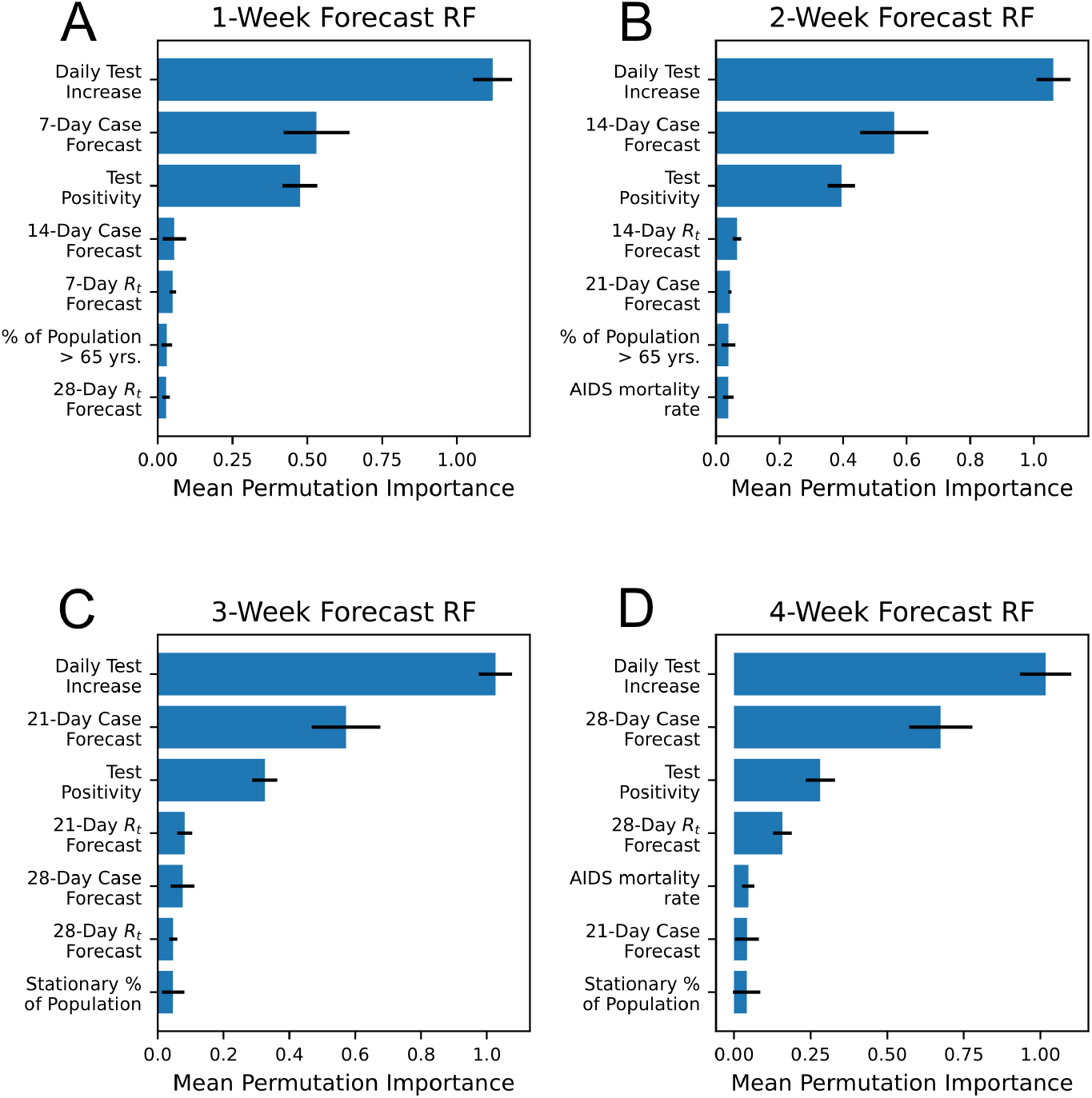
Top 7 RF Features for Each Forecast Target. For each forecast target, RF feature permutation importances were averaged over all 11 epi weeks and the top 7 features are shown in the subfigures above along with their standard deviation as error bars.

On the training datasets for all 4 forecast targets and all 11 epi weeks, *R*^2^ was very stable, averaging 0.97 with a standard deviation of 0.00. For the corresponding validation datasets, *R*^2^ fell to 0.92 with a standard deviation of 0.02. Thus, overfitting is not extreme and, considering the RFs’ relatively high MAE and *R*^2^ on actual case data in comparison to other models as seen in Fig. 4-5, is not detrimental to our objectives.

**Figure 4:**
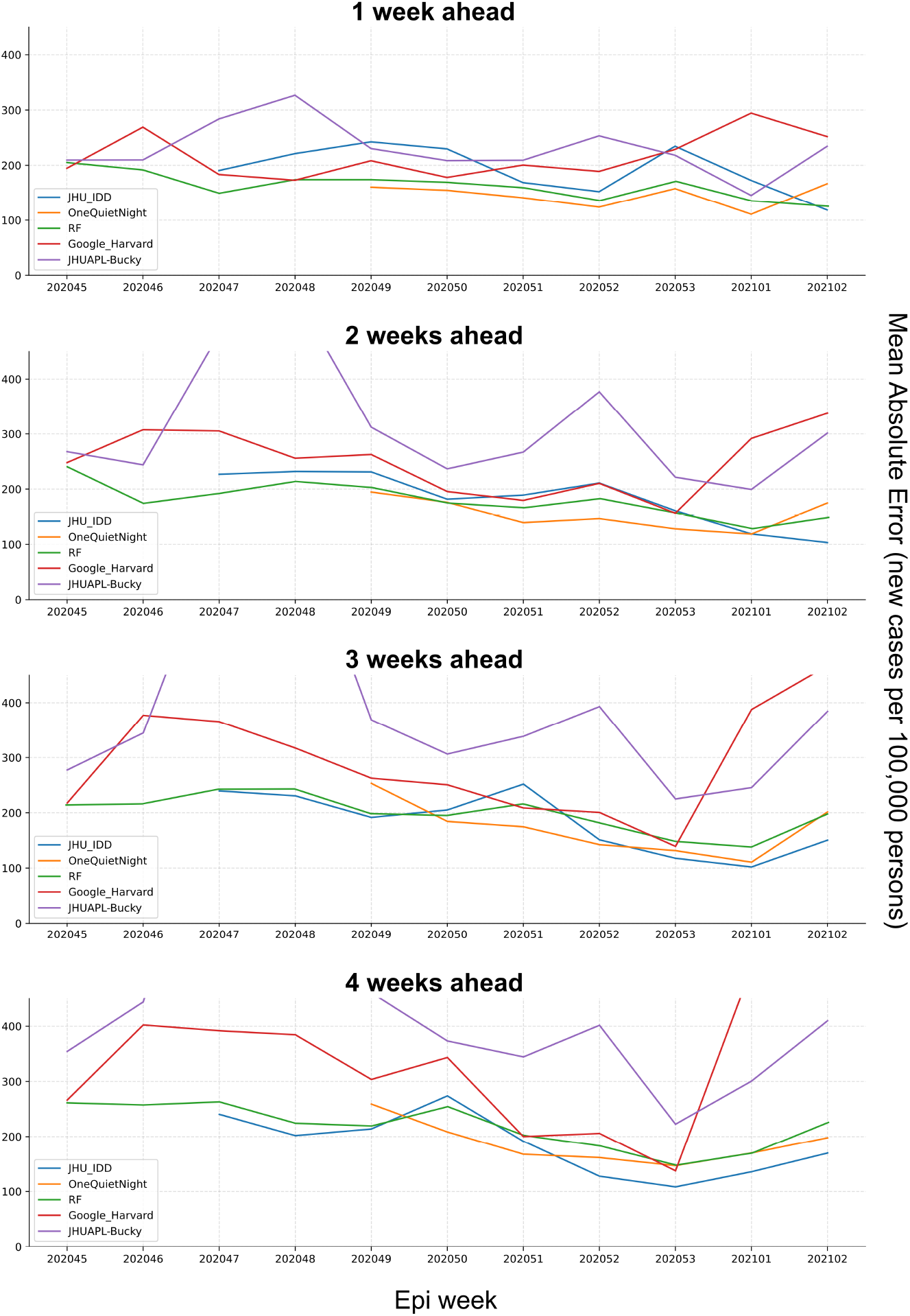
Performance Evaluation Using MAE. The errors between projections and real number of incident cases were calculated using Eq. 3. The y-axes of the graphs have been limited so that all models can be visually compared.

**Figure 5:**
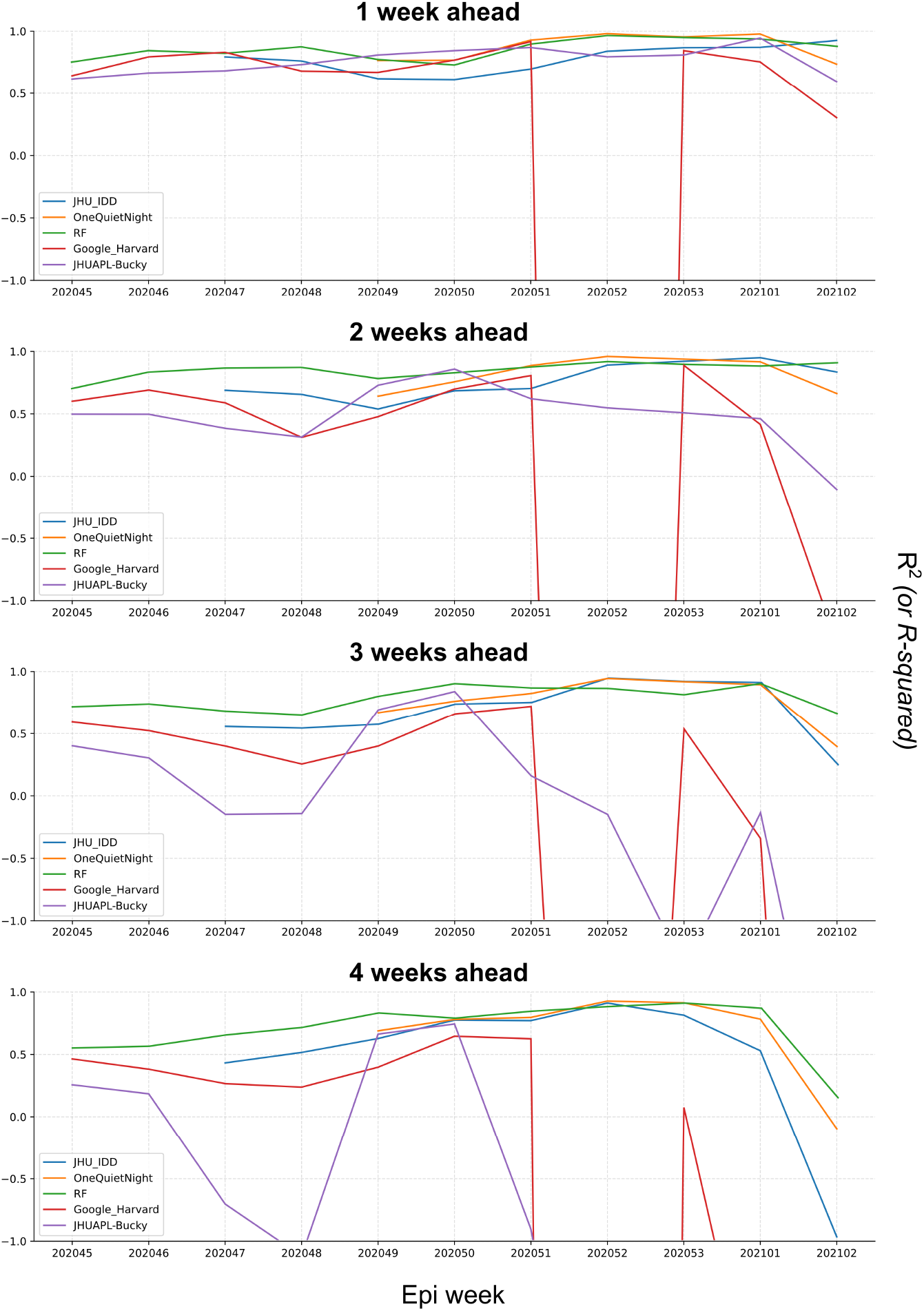
Performance Evaluation Using R^2^. As with Fig. 4, the y-axes of the graphs have been limited. The proportions of variance between projections and observed values were evaluated using Eq. 2. We notice that there are large anomalies in the weekly *R*^2^ of the Google_Harvard and JHUAPL_Bucky models after epi week 202051; however, for the sake of complete comparison, all weeks for all models are shown.

### 3.3 RF Forecasting Performance vs. Other Models

As may be observed in Fig. 4 and Fig.5, over the 11 reference weeks for which forecasts were generated by the RF models, MAE and *R*^2^ remain relatively consistent compared to the OQN and JHU models. The RF’s *R*^2^ is competitive with and often higher than the *R*^2^ for other models, notably November 2020. Cases in November were harder to model accurately, as November saw a 40% increase in cases in the 3068 counties in its fourth week relative to its first week, whereas December only saw an 8% decrease in cases for its equivalent comparison period. Periods of relatively low performance are generally shared by all models, indicating that case load changes during these weeks are simply less predictable.

For each epi week, case projections from each model were collected and compared to actual cases that occurred during these weeks as reported by JHU [18]. The results were calculated and visualized in Fig. 4-5.

The JHU IDD and APL models are SEIR models [29, 30], whereas the OQN model applies a linear regression model to each CCE [31]. The distribution of output from a model exclusive to a CCE will be skewed towards the distribution of its training dataset labels, which may be a factor explaining OQN’s low MAE. On the other hand, the RF model forecasts for all counties, affording it a larger dataset which possibly contributes to its relatively high, stable fit to the actual cases as indicated by its *R*^2^.

## 4 Conclusion

We present a unique method to project COVID-19 cases for CCEs by using their or their state’s *R*_*t*_ time-series to predict cases, taking advantage of the backward lag of regional *R*_*t*_ time-series from the case time-series despite their similar trajectory. These predictions are input into a RF regression models with regional testing, demographic, population mobility and population health data for final case forecasts. Our approach is computationally inexpensive while remaining very effective, as our model achieves consistently high *R*^2^ and low MAE relative to gold standard models used in the U.S. CDC ensemble model during a highly dynamic case spike period in November 2020 – early January 2021. This model is limited to forecasting cases detected by testing as opposed to latent, asymptomatic cases, which can be estimated by compartmental models [35]. Thus, its best use scenario is for public health officials to identify potential outbreaks in their community to help them optimize their response. It shows evidence of good consistency in its current iteration but has some potential for improvement via addition of new features to its training dataset, particularly case and *R*_*t*_ forecasts from other compartmental models. Such ensemble forecasting approaches have improved model performance significantly [36].

## Data Availability

All code and program supporting the conclusions made in this article is publicly available on GitHub. Our repository can be found at https://github.com/solveforj/pandemic-central. Our weekly forecasts are updated and visualized at https://itsonit.com.

## 5 Supplementary Information

### 5.1 Acknowledgements

We would like to thank Dr. Richard P. Olenick (University of Dallas) for providing us with insightful criticism of the manuscript. We also wish to thank all organizations that curate our input datasets and make them freely available for researchers.

### 5.2 Availability of software and other materials

All code and program supporting the conclusions made in this article is publicly available on GitHub. Our repository can be found at https://github.com/solveforj/pandemic-central.

Our weekly forecasts are updated and visualized on Pandemic Central, available at https://itsonit.com.

### 5.3 Declaration of Competing Interest

We have read and fully understood the competing interest policy. Projections from multiple versions of our model are voluntarily uploaded to the COVID-19 Forecast Hub since August 2020 and were published by the U.S. CDC [37]. We wish to declare that:

- We have no competing interests or external funding.
- We do not receive any form of support, including financially, from the U.S. CDC or COVID-19 Forecast Hub.
- We do not have any personal association with organizations in prior 36 months that may have any effect on the outcome of this research.

## Notes

### Competing Interest Statement

The authors have declared no competing interest.

### Funding Statement

No external funding was received to support this study.

## References

[1] T. Walmsley, A. Rose, and D. Wei, “The impacts of the coronavirus on the economy of the united states,” SSRN Electronic Journal, 2020. doi: 10.2139/ssrn.3678835

[2] C. W. Yancy, “COVID-19 and african americans,” JAMA, vol. 323, no. 19, p. 1891, May 2020. doi: 10.1001/jama.2020.6548

[3] L. Bertozzi, E. Franco, G. Mohler, M. B. Short, and D. Sledge, “The challenges of modeling and forecasting the spread of COVID-19,” Proceedings of the National Academy of Sciences, vol. 117, no. 29, pp. 16 732–16 738, Jul 2020. doi: 10.1073/pnas.2006520117

[4] W. Messner and S. Payson, “Variation in COVID-19 outbreaks at the US state and county levels,” Public Health, vol. 187, pp. 15–18, Oct 2020. doi: 10.1016/j.puhe.2020.07.035. [Online]. Available: https://doi.org/10.1016%2Fj.puhe.2020.07.035

[5] H. M. Paiva, R. J. M. Afonso, I. L. de Oliveira, and G. F. Garcia, “A data-driven model to describe and forecast the dynamics of COVID-19 transmission,” PLOS ONE, vol. 15, no. 7, p. e0236386, Jul 2020. doi: 10.1371/journal.pone.0236386

[6] S. R. Buckman, R. Glick, K. J. Lansing, N. Petrosky-Nadeau, and L. M. Seitelman, “Replicating and projecting the path of COVID-19 with a model-implied reproduction number,” Infectious Disease Modelling, vol. 5, pp. 635–651, 2020. doi: 10.1016/j.idm.2020.08.007

[7] Özgür Özmen, J. J. Nutaro, L. L. Pullum, and A. Ramanathan, “Analyzing the impact of modeling choices and assumptions in compartmental epidemiological models,” SIMULATION, vol. 92, no. 5, pp. 459–472, Apr 2016. doi: 10.1177/0037549716640877

[8] Zeroual, F. Harrou, A. Dairi, and Y. Sun, “Deep learning methods for forecasting COVID-19 time-series data: A comparative study,” Chaos, Solitons & Fractals, vol. 140, p. 110121, Nov 2020. doi: 10.1016/j.chaos.2020.110121

[9] C. Shang,, K. E. Galow, G. G. Galow, and and, “Regional forecasting of COVID-19 caseload by nonparametric regression: a VAR epidemiological model,” AIMS Public Health, vol. 8, no. 1, pp. 124–136, 2021. doi: 10.3934/publichealth.2021010

[10] X. Fang, W. Liu, J. Ai, M. He, Y. Wu, Y. Shi, W. Shen, and C. Bao, “Forecasting incidence of infectious diarrhea using random forest in jiangsu province, china,” BMC Infectious Diseases, vol. 20, no. 1, Mar 2020. doi: 10.1186/s12879-020-4930-2

[11] M. H. D. M. Ribeiro, R. G. da Silva, V. C. Mariani, and L. dos Santos Coelho, “Short-term forecasting COVID-19 cumulative confirmed cases: Perspectives for brazil,” Chaos, Solitons & Fractals, vol. 135, p. 109853, Jun 2020. doi: 10.1016/j.chaos.2020.109853

[12] L. Breiman, “Random forests,” Machine Learning, vol. 45, no. 1, p. 5–32, Oct 2001. doi: 10.1023/A:1010933404324

[13] X. Chen and H. Ishwaran, “Random forests for genomic data analysis,” Genomics, vol. 99, no. 6, pp. 323–329, Jun 2012. doi: 10.1016/j.ygeno.2012.04.003

[14] G. Machado, M. R. Mendoza, and L. G. Corbellini, “What variables are important in predicting bovine viral diarrhea virus? a random forest approach,” Veterinary Research, vol. 46, no. 1, Jul 2015. doi: 10.1186/s13567-015-0219-7

[15] E. Mussumeci and F. C. Coelho, “Large-scale multivariate forecasting models for dengue - LSTM versus random forest regression,” Spatial and Spatio-temporal Epidemiology, vol. 35, p. 100372, Nov 2020. doi: 10.1016/j.sste.2020.100372

[16] M. J. Kane, N. Price, M. Scotch, and P. Rabinowitz, “Comparison of ARIMA and random forest time series models for prediction of avian influenza h5n1 outbreaks,” BMC Bioinformatics, vol. 15, no. 1, Aug 2014. doi: 10.1186/1471-2105-15-276

[17] C. Keyel, O. E. Timm, P. B. Backenson, C. Prussing, S. Quinones, K. A. McDonough, M. Vuille, J. E. Conn, P. M. Armstrong, T. G. Andreadis, and L. D. Kramer, “Seasonal temperatures and hydrological conditions improve the prediction of west nile virus infection rates in culex mosquitoes and human case counts in new york and connecticut,” PLOS ONE, vol. 14, no. 6, p. e0217854, Jun 2019. doi: 10.1371/journal.pone.0217854

[18] “COVID-19 map,” Available: https://coronavirus.jhu.edu/map.html.

[19] “Movement range maps,” Available: https://data.humdata.org/dataset/movement-range-maps.

[20] “The COVID tracking project,” Available: https://covidtracking.com.

[21] “COVID Act Now,” Available: https://covidactnow.org/data-api.

[22] “United states infectious disease mortality rates by county 1980-2014,” Available: http://ghdx.healthdata.org/record/ihme-data/united-states-infectious-disease-mortality-rates-county-1980-2014.

[23] “United states chronic respiratory disease mortality rates by county 1980-2014,” Available: http://ghdx.healthdata.org/record/ihme-data/united-states-chronic-respiratory-disease-mortality-rates-county-1980-2014.

[24] “United states life expectancy and age-specific mortality risk by county 1980-2014,” Available: http://ghdx.healthdata.org/record/ihme-data/united-states-life-expectancy-and-age-specific-mortality-risk-county-1980-2014.

[25] “United states diabetes prevalence by county 1999-2012,” Available: http://ghdx.healthdata.org/record/ihme-data/united-states-diabetes-prevalence-county-1999-2012.

[26] “United states physical activity and obesity prevalence by county 2001-2011,” Available: https://doi.org/10.6069/5E84-HD26.

[27] “County population by characteristics,” Available: https://www.census.gov/data/datasets/time-series/demo/popest/2010s-counties-detail.html.

[28] F. Pedregosa, G. Varoquaux, A. Gramfort, V. Michel, B. Thirion, O. Grisel, M. Blondel, P. Prettenhofer, R. Weiss, V. Dubourg, J. Vanderplas, A. Passos, D. Cournapeau, M. Brucher, M. Perrot, and E. Duchesnay, “Scikit-learn: Machine learning in python,” Journal of Machine Learning Research, vol. 12, no. 85, Oct 2011. [Online]. Available: https://jmlr.org/papers/volume12/pedregosa11a/pedregosa11a.pdf

[29] J. C. Lemaitre, K. H. Grantz, J. Kaminsky, H. R. Meredith, S. A. Truelove, S. A. Lauer, L. T. Keegan, S. Shah, J. Wills, K. Kaminsky, J. Perez-Saez, J. Lessler, and E. C. Lee, “A scenario modeling pipeline for covid-19 emergency planning,” Scientific Reports, vol. 11, no. 1, p. 7534, Apr 2021. doi: 10.1038/s41598-021-86811-0

[30] The Johns Hopkins University Applied Physics Laboratory, “Bucky’s documentation,” 2020, unpublished. [Online]. Available: https://docs.buckymodel.com/en/latest/index.html

[31] Jo and J. Cho, “OneQuietNight COVID-19 forecast,” 2021, unpublished manuscript. [Online]. Available: https://one-quiet-night.github.io/vis/static/media/OQN.631fe207.pdf

[32] E. L. Ray, N. Wattanachit, J. Niemi, A. H. Kanji, K. House, E. Y. Cramer, J. Bracher, A. Zheng, T. K. Yamana, X. Xiong, S. Woody, Y. Wang, L. Wang, R. L. Walraven, V. Tomar, K. Sherratt, D. Sheldon, R. C. Reiner, B. A. Prakash, D. Osthus, M. L. Li, E. C. Lee, U. Koyluoglu, P. Keskinocak, Y. Gu, Q. Gu, G. E. George, G. España, S. Corsetti, J. Chhatwal, S. Cavany, H. Biegel, M. Ben-Nun, J. Walker, R. Slayton, V. Lopez, M. Biggerstaff, M. A. Johansson, and N. G. Reich, “Ensemble forecasts of coronavirus disease 2019 (COVID-19) in the u.s.” 2020, unpublished manuscript. [Online]. Available: https://www.medrxiv.org/content/early/2020/08/22/2020.08.19.20177493

[33] “Clinical questions about COVID-19: Questions and answers,” Available: https://www.cdc.gov/coronavirus/2019-ncov/hcp/faq.html.

[34] R. Omori, K. Mizumoto, and G. Chowell, “Changes in testing rates could mask the novel coronavirus disease (COVID-19) growth rate,” International Journal of Infectious Diseases, vol. 94, pp. 116–118, may 2020. doi: 10.1016/j.ijid.2020.04.021

[35] Ra?dulescu, C. Williams, and K. Cavanagh, “Management strategies in a SEIR-type model of COVID-19 community spread,” Scientific Reports, vol. 10, no. 1, dec 2020. doi: 10.1038/s41598-020-77628-4

[36] N. E. Dean, A. P. y Piontti, Z. J. Madewell, D. A. Cummings, M. D. Hitchings, K. Joshi, R. Kahn, A. Vespignani, M. E. Halloran, and I. M. Longini, “Ensemble forecast modeling for the design of COVID-19 vaccine efficacy trials,” Vaccine, vol. 38, no. 46, pp. 7213–7216, oct 2020. doi: 10.1016/j.vaccine.2020.09.031

[37] “COVID-19 forecast hub,” Available: https://github.com/reichlab/covid19-forecast-hub.

